# Distribution of age at natural menopause, age at menarche, menstrual cycle length, height and BMI in BRCA1 and BRCA2 pathogenic variant carriers and non-carriers: Results from EMBRACE

**DOI:** 10.1101/2025.02.12.25321672

**Authors:** Nasim Mavaddat, Debra Frost, Emily Zhao, Daniel R Barnes, Munaza Ahmed, Julian Barwell, Angela F Brady, Paul Brennan, Hector Conti, Jackie Cook, Harriet Copeland, Rosemarie Davidson, Alan Donaldson, Emma Douglas, David Gallagher, Rachel Hart, Louise Izatt, Zoe Kemp, Fiona Lalloo, Zosia Miedzybrodzka, Patrick J Morrison, Jennie E. Murray, Alex Murray, Hannah Musgrave, Claire Searle, Lucy Side, Katie Snape, Vishakha Tripathi, Lisa Walker, Stephanie Archer, D. Gareth Evans, Marc Tischkowitz, Antonis C Antoniou, Douglas F. Easton

## Abstract

**Background:** Carriers of germline pathogenic variants (PVs) in the BRCA1 and BRCA2 genes are at higher risk of developing breast and ovarian cancer than the general population. It is unclear if these PVs influence other breast or ovarian cancer risk factors, including age at menopause (ANM), age at menarche (AAM), menstrual cycle length, BMI or height. There is a biological rationale for associations between BRCA1 and BRCA2 PVs and reproductive and anthropomorphic traits, for example involving DNA damage and repair mechanisms. The evidence for or against such associations is limited.

**Methods:** We used data on 3,046 BRCA1 and 3,264 BRCA2 PV carriers, and 2,857 non-carrier female relatives of PV carriers from the Epidemiological Study of Familial Breast Cancer (EMBRACE). Associations between ANM and PV carrier status was evaluated using linear and Cox regression models allowing for censoring. AAM, menstrual cycle length, BMI, and height in carriers and non-carriers were compared using linear and multinomial logistic regression. Analyses were adjusted for potential confounders, and weighted analyses carried out to account for non-random sampling with respect to cancer status.

**Results:** No statistically significant difference in ANM between carriers and non-carriers was observed in analyses accounting for censoring. Linear regression effect sizes for ANM were - -0.002 (95%CI: -0.401, 0.397) and -0.172 (95%CI: -0.531, 0.188), for BRCA1 and BRCA2 PV carriers respectively, compared with non-carrier women. The distributions of AAM, menstrual cycle length and BMI were similar between PV carriers and non-carriers, but BRCA1 PV carriers were slightly taller on average than non-carriers (0.5cm difference, p=0.003).

**Conclusion:** Contrary to previous reports, we found no evidence that BRCA1 or BRCA2 PV are associated with hormonal or anthropometric factors, except for a weak association with height. These results inform the incorporation of risk factors into multifactorial cancer risk prediction algorithms.

## Introduction

Germline pathogenic variants in BRCA1 and BRCA2 confer high risks of breast and ovarian cancer (1). Reproductive factors, including age at natural menopause (ANM) and age at menarche (AAM), and anthropomorphic traits including height and body mass index (BMI) are established breast and/or ovarian cancer risk factors in the general population (2). There is evidence from observational studies and Mendelian randomisation analyses that some breast cancer risk factors in the general population are also associated with cancer risk in PV carriers (3–9). Risk prediction algorithms, notably BOADICEA, incorporate the effects of both PVs and other risk factors to predict cancer risk (10, 11). These algorithms depend on assumptions about the distribution of these traits in PV carriers, and well as the associated effect sizes. Further, management of carriers may include recommendation on risk reducing bilateral salpingo-oophorectomy (RRSO) and the likely timing of menopause may be an important consideration for women contemplating surgery.

Age at natural menopause is normally distributed in the general population with an average age of ∼50 years in European ancestry women. Menopause occurs between ages 40-60 years in 99% of women and before age 40 years in ∼1% of women; women with age at menopause less than 40 years may be diagnosed with premature ovarian insufficiency, a largely monogenic trait. Certain environmental factors are associated with earlier ANM, including lower BMI, alcohol, smoking and low birth weight. Maternal obesogenic diet during pregnancy also decreases the ovarian reserve in offspring (12, 13). ANM has a strong genetic basis, mediated by multiple genetic loci, many of which have been identified through genome-wide association analyses (13). ANM associated SNPs are enriched for variants near genes involved in DNA damage response. These include common coding variants in BRCA1: the alleles associated with earlier ANM are also associated with reduced BRCA1 expression in blood (14). These common variants have not, however, been associated with cancer risk (15). BRCA1 expression decreases in human ovaries with age (16), while reduced brca1 expression in mouse models leads to reduced ovarian reserve (16). BRCA1 directly inhibits a functional interaction with oestrogen receptor α and thus BRCA1 variants could also affect ANM through altered oestrogen signalling (17).

In exome-wide analysis in UK Biobank data, rare loss-of-function (LOF) variants in BRCA1 and BRCA2 were associated with earlier (2.63 and 1.53 years respectively) ANM compared with non-carriers, while LOF variants in CHEK2 were associated later ANM (3.49 years difference) (13). Rare coding variants in other DNA damage repair genes have also been associated with ANM (18). Earlier epidemiological studies have suggested that natural menopause occurs at a younger age in BRCA1 and BRCA2 PV carriers compared with women from the general population (19–21), and that BRCA1 PV carriers may have reduced ovarian reserve (22) and consequently a shortened reproductive lifespan. Other studies, however, have reported no statistically significant differences between ANM in BRCA1/2 carriers and the general population (23). These analyses are, however, complicated by incompleteness of data on preventative surgeries, in particular RRSO, and potential reverse causation as a diagnosis of cancer and associated treatments may also be associated with onset of menopause. Age at menarche, weight and height are also highly heritable polygenic traits, with both rare variant and polygenic influences (24–26). Many of the AAM-linked genetic variants discovered in the general population (27) are also associated with BMI.

A number of studies have investigated association between reproductive and anthropomorphic traits and cancer risk in BRCA1 and BRCA2 PV carriers (28–30), but few studies have evaluated their distribution in comparable carrier and non-carrier populations. Here we used data from the Epidemiological Study of Familial Breast Cancer (EMBRACE), a large national study of PV carriers and non-carrier relatives, to evaluate differences in reproductive and anthropometric trait distributions among BRCA1 and BRCA2 PV carriers and non-carriers. These data can be used to adapt risk prediction algorithms for PV carriers and may further inform our understanding of reproductive biology of female carriers of PVs in these susceptibility genes.

## Methods

### Study design and population

Participants were enrolled through an on-going nationwide study of individuals undergoing genetic testing in regional genomics centres in the United Kingdom and Ireland (EMBRACE) (https://ccge.medschl.cam.ac.uk/embrace/). EMBRACE recruits individuals who are carriers of pathogenic variants (PVs) in breast and/or ovarian cancer susceptibility genes, and their relatives. The analysis reported here included only women of self-reported White ethnicity. Women were eligible if they were at least 18 years of age at recruitment and had tested positive for a BRCA1 or BRCA2 PV or were non-carrier family members of PV carriers. PVs were defined according to ENIGMA/ClinGen guidelines (https://clinicalgenome.org/affiliation/50087/).

### Data collection

All study participants were invited to complete a baseline questionnaire requesting detailed information on known or suspected risk factors for breast and ovarian cancer, including family history of cancer, height, weight at age 18, current weight, reproductive history and surgical interventions including risk-reducing mastectomy (RRM) or RRSO. The questionnaires also requested information on age at last menstruation, whether the woman had had any period in the past year, the number of years/months since last menstruation, and reason(s) for periods stopping. PV carriers also completed follow-up questionnaires: however, since these were not completed by non-carriers and the primary interest was the comparison of carriers and non-carriers, only information from the baseline questionnaire was used here.

Women were considered premenopausal if they indicated that they had had a period in the past year, or if the ‘reason for periods stopping’ was medication or oral contraceptive use (unless 40 years or older), pregnancy or breast-feeding. Age at menopause for those who indicated no period in the past year was determined by adding 11year to ‘age at last menstruation’. Women were considered as having experienced natural menopause if the reason for periods stopping was recorded as ‘natural menopause’ (and not for any other reason such as chemotherapy, childbirth, pregnancy, breast feeding or hysterectomy) and age at menopause preceded RRSO, any cancer diagnosis (apart from non-melanoma skin cancer), or interview. Women were also considered as menopausal at age 55 years. Women reporting RRSO or hysterectomy as the reason for periods stopping were considered premenopausal until the age at last period. Women reporting periods stopping (due to natural menopause, RRSO or hysterectomy) but with missing age at menopause or age at last period were excluded from the analyses.

The numbers of women experiencing RRSO at censoring are shown in STable 1 and the reasons for censoring by menopausal status are summarised in STable 2. The numbers of breast cancers diagnosed prior to or at interview by age at diagnosis are shown in STable 3.

Age at menarche was coded as a continuous variable or categorised as age <12, 12-14 and ≥15 years. The interval between menarche and the earliest of menopause and age at censoring (years) was treated as a continuous variable. Women were asked if occurrence of menstrual cycle was always regular, usually regular, or never regular. For women with always regular or usually regular cycles, menstrual cycle length information was categorised as <26, 26-27 and ≥28 days. Parity at baseline was coded as nulliparous, one, two, or three or more live births. Age at first birth was categorised as a continuous or categorical variable (age <20, 21-25, 26-30, 31-35, ≥35 years). Height (m) was treated as a continuous variable. BMI was calculated as weight (kg) divided by height (m) squared.

### Statistical Analyses

We carried out three types of analysis to explore whether age at natural menopause was influenced by PV carrier status. The association between carrier status and ANM was first explored using linear regression, including only women experiencing natural menopause. These analyses were adjusted for birth cohort (as described below), and age-group at censoring (in two-year categories from <40 to ≥ 54 years), the last age at which menopause could be observed. We also carried out linear regression analyses allowing for a censored outcome, using the cens.normal function in the VGAM package in R (https://CRAN.R-project.org/package=VGAM and (31, 32)). Women were censored at the earliest of age at natural menopause, age at RRSO, any cancer diagnosis apart from non-melanoma skin cancer, death, age at interview or age 55 years. This analysis allowed pre-menopausal women (right censored at baseline) as well as post-menopausal women to be included but assumes that carrier status shifts the mean ANM (rather than the proportional hazards assumption made in a Cox regression). These analyses were also used to evaluate the association between carrier status and the interval between ANM and AAM, and carrier status.

Finally, time-to-event analyses were carried out using Cox regression, with entry starting at birth and followed-up until the earliest of age at menopause, age at RRSO, any cancer diagnosis, death, age at interview or age 55 years. However, it should be noted that treating the data as arising from a “true” cohort is not strictly valid, since individuals were recruited at different ages as adults and the data considered here are retrospective, not prospective.

Linear regression models were used to test for associations between PV carrier status and AAM, menstrual cycle length, height, BMI at interview and BMI at age 18 years. Associations with categorical AAM and menstrual cycle length was also assessed using multinomial logistic regression.

Participants in EMBRACE were recruited from a population undergoing genetic testing. Affected individuals are therefore more likely to be sampled than unaffected individuals. Additionally, there is a higher probability of sampling younger affected individuals. To account for this bias, a weighted cohort method in which affected and unaffected women are assigned different weights in all analyses according to their age at diagnosis, or age at censoring, was used so that the weighted cohort mimicked a true cohort (33, 34). This method has been shown to provide estimates of relative risk which are close to unbiased (33, 34). An individual was considered a case if they had had a breast cancer diagnosis prior to or at age at interview, regardless of menopausal status, and otherwise a control. For calculation of weights the person-years for unaffected women were calculated from birth to the first of age at interview or RRM, while the person-years for affected women were from birth to age at breast cancer diagnosis, regardless of menopausal status. Individuals were weighted such that the observed breast cancer incidence rates were consistent with established age-specific incidence rates and relative risk estimates for BRCA1 and BRCA2 PV carriers (11, 35, 36) (STables 3 and 4). Non-carriers were not weighted (weight=1) as the proportion of non-carriers that were affected was small (37).

Analyses of ANM were adjusted by birth cohort (year of birth <1940, 1940-1949,1950-1959 and ≥1960); by parity, with the number of full-term pregnancies categorised as 0, 1, 2, and 3 or more; and by age at the start of first full-term pregnancy, categorised as <20, 20-24, 25-29, 30-34 and ≥35 years. Analyses were carried out clustering for family membership, and robust variance-adjusted confidence intervals reported.

For analyses of AAM, menstrual cycle length, height, BMI at interview and BMI at age 18 years, models were adjusted using a finer categorisation of birth cohort (i.e. splitting the final category into 1960-1969 and ≥1970 groups). For AAM, analyses were also adjusted for BMI at age 18 years. When evaluating menstrual cycle length, analyses were also adjusted for age at interview.

All statistical analyses were conducted using R version 4.3.1 and associated packages.

## Results

### Study participants

A total of 3,046 BRCA1 PV carriers, 3,264 BRCA2 PV carriers and 2,857 non-carriers from EMBRACE were included in the analyses. Cohort characteristics and distribution of reproductive risk factors, height and BMI are shown in STable 1. The distribution of age at interview was similar between carriers and non-carriers. Approximately 44% of carriers had been diagnosed with breast cancer at interview, compared with ∼3.6% of non-carriers (STable 3).

### Distribution of age at natural menopause among BRCA1 and BRCA2 carriers and non-carriers

Among women included in the analysis, 379 (12%) of BRCA1 PV carriers, 646 (20%) of BRCA2 PV carriers and 645 (23%) of non-carriers experienced natural menopause prior to RRSO, a cancer diagnosis (apart from non-melanoma skin cancer) or interview (STable 1).

The mean ANM was lower among BRCA1 carriers than non-carriers (50.0 vs 50.8 years respectively, STable 1) and this difference was statistically significant in linear regression analysis unadjusted for age at censoring (p=0.01, Table 1). However, this difference was no longer apparent when analyses were adjusted for the age at censoring, the last age at which menopause could have been observed (ANM carrier vs. non-carrier difference -0.129 years, (95%CI: -0.578, 0.321)). There was also no effect of carrier status on ANM in linear regression analyses allowing for a censored outcome, which included data from both pre- and post-menopausal women (ANM difference=-0.002 (95%CI: -0.401, 0.397)).

**Table 1:** Association between age at natural menopause and BRCA1 and BRCA2 PV carrier vs non-carrier status.

|  | ***BRCA1* PV carriers** |  |  |  | ***BRCA2* PV carriers** |  |  |  |
| --- | --- | --- | --- | --- | --- | --- | --- | --- |
|  | estimate | L95CI | U95CI | p-value | estimate | L95CI | U95CI | p-value |
| Linear regression among menopausal women only |  |  |  |  |  |  |  |  |
| + Birth cohort | -0.700 | -1.241 | -0.159 | 0.011 | -0.215 | -0.686 | 0.255 | 0.369 |
| + Cens agegroup | -0.129 | -0.578 | 0.321 | 0.574 | -0.171 | -0.602 | 0.260 | 0.437 |
| + Cens agegroup+Parity+AFB+BMI18+AAM<sup>a</sup> | -0.113 | -0.570 | 0.343 | 0.625 | -0.074 | -0.509 | 0.361 | 0.739 |
| + Cens agegroup+Parity+AFB<sup>b</sup> | -0.098 | -0.579 | 0.383 | 0.688 | -0.058 | -0.510 | 0.393 | 0.800 |
| Linear regression among pre and post-menopausal women using cens.normal function in VGAM |  |  |  |  |  |  |  |  |
| + Birth cohort | -0.002 | -0.401 | 0.397 | 0.991 | -0.172 | -0.531 | 0.188 | 0.349 |
| + Parity+AFB+BMI+AAM<sup>a</sup> | -0.052 | -0.451 | 0.347 | 0.798 | -0.139 | -0.500 | 0.222 | 0.451 |
| + Parity+AFB+BMI18+AAM<sup>a</sup> | 0.006 | -0.395 | 0.406 | 0.977 | -0.104 | -0.467 | 0.260 | 0.576 |
| In women with ANM or end of FUP >=40 years | 0.077 | -0.302 | 0.457 | 0.69 | 0.002 | -0.334 | 0.339 | 0.989 |

ANM, age at natural menopause; AAM, age at menarche; AFB, age at first birth; BMI, Body Mass Index at baseline; BMI18, BMI at age 18 years; 'Cens agegroup', refers to analyses adjusted for the last age at which menopause could be observed; FUP, follow-up Analyses were adjusted for birth cohort (categorised as <1940, 1940-1949, 1950-1959, >=1960) and using weights derived as described in the Methods.
<sup>a</sup> carried out on data with no missing information on AAM, BMI or BMI at age 18, parity; AFB and parity were considered as categorical variables
<sup>b</sup> only among parous women with information on AFB; AFB was considered as a continuous variable

Similarly, there was no difference in distribution of age at menopause between BRCA2 carriers and non-carriers (mean age at menopause among BRCA2 PV carriers= 50.6 years; linear regression coefficient=-0.172 (95%CI: -0.531, 0.188), in analyses allowing for a censored outcome) (Table 1). Adjustment for BMI parity, and age at first birth did not materially alter the estimates.

In the time-to-event analyses using Cox regression, HRs were 0.99, (95%CI: 0.88-1.11) and 0.99 (95%CI:0.90-1.10) for BRCA1 and BRCA2 PV carriers respectively, p>0.05 (STable 5) in unadjusted analyses. Models adjusting for age at menarche, BMI at age 18, parity and age at first birth gave similar results (STable 5).

### Distribution of age at menarche, menstrual cycle length and reproductive lifespan among BRCA1 and BRCA2 carriers and non-carriers

Mean age at menarche was 12.97 and 12.90 years for BRCA1 and BRCA2 carriers respectively, and 12.94 years among non-carriers. There was no statistically significant difference in age at menarche either alone (as a continuous or categorical variable) or after adjusting for BMI (Table 2). There were no statistically significant differences between carriers and non-carriers in menstrual cycle length (in women with always regular or usually regular cycles) (Table 3). The interval between menarche and age at menopause was also similar between carriers and non-carriers in regression analyses allowing for censoring (Table 4).

**Table 2:** Association between age at menarache and BRCA1 and BRCA2 PV carrier vs non-carrier status.

|  | BRCA1 PV carriers |  |  |  | BRCA2 PV carriers |  |  |  |
| --- | --- | --- | --- | --- | --- | --- | --- | --- |
| Linear Regression |  |  |  |  |  |  |  |  |
|  | estimate | L95CI | U95CI | p-value | estimate | L95CI | U95CI | p-value |
| AAM as continuous variable +Birth cohort | 0.068 | -0.014 | 0.150 | 0.106 | -0.071 | -0.152 | 0.010 | 0.084 |
| AAM as categorical variable - trend +Birth cohort | 0.027 | -0.004 | 0.058 | 0.085 | -0.021 | -0.052 | 0.010 | 0.187 |
| Analyses among women with no missing information on AAM, BMI, and height |  |  |  |  |  |  |  |  |
| +Birth cohort | 0.064 | -0.019 | 0.147 | 0.130 | -0.076 | -0.158 | 0.005 | 0.067 |
| +Birth cohort+BMI18 | 0.064 | -0.018 | 0.146 | 0.127 | -0.071 | -0.152 | 0.010 | 0.085 |
| +Birth cohort+BMI18+height | 0.056 | -0.027 | 0.138 | 0.186 | -0.075 | -0.156 | 0.006 | 0.069 |
| Multinomial regression (+Birth cohort) |  |  |  |  |  |  |  |  |
|  | OR | L95CI | U95CI | p-value | OR | L95CI | U95CI | p-value |
| <12 years | 1.000 |  |  |  | 1.000 |  |  |  |
| 12-14 years | 1.115 | 0.974 | 1.276 | 0.115 | 0.940 | 0.826 | 1.069 | 0.344 |
| >=15 years | 1.171 | 0.980 | 1.400 | 0.082 | 0.884 | 0.743 | 1.053 | 0.167 |

AAM, Age at menarche; BMI, Body Mass Index at baseline; BMI18, BMI at age 18 years.
Analyses were adjusted for birth cohort (categorised as <1940, 1940-1949, 1950-1959, 1960-1969, 1970-1979, >=1980); and using weights derived as described in the Methods.

**Table 3:** Association between menstrual cycle length and BRCA1 and BRCA2 PV carrier vs non-carrier status.

|  | BRCA1 PV carriers |  |  |  | BRCA2 PV carriers |  |  |  |
| --- | --- | --- | --- | --- | --- | --- | --- | --- |
| Analyses among women with no missing information on AAM, and with always regular or usually regular periods |  |  |  |  |  |  |  |  |
| Linear Regression (+Birth cohort) |  |  |  |  |  |  |  |  |
|  | estimate | L95CI | U95CI | p-value | estimate | L95CI | U95CI | p-value |
| Menstrual cycle length (days) | 0.139 | -0.017 | 0.295 | 0.081 | 0.030 | -0.115 | 0.175 | 0.688 |
| Multinomial regression (+Birth cohort) |  |  |  |  |  |  |  |  |
|  | OR | L95CI | U95CI | p-value | OR | L95CI | U95CI | p-value |
| <26 years | 1.000 |  |  |  | 1.000 |  |  |  |
| 26-27 | 1.006 | 0.793 | 1.278 | 0.958 | 1.131 | 0.897 | 1.426 | 0.297 |
| >=28 | 1.041 | 0.883 | 1.229 | 0.630 | 1.045 | 0.887 | 1.231 | 0.597 |
| Analyses among women with no missing information on AAM, BMI, height and parity and always regular or usually regular periods |  |  |  |  |  |  |  |  |
| Linear Regression (+Birth cohort) |  |  |  |  |  |  |  |  |
|  | estimate | L95CI | U95CI | p-value | estimate | L95CI | U95CI | p-value |
| Menstrual cycle length (days) | 0.143 | -0.012 | 0.299 | 0.071 | 0.029 | -0.117 | 0.174 | 0.699 |
| +AAM+height+BMI+Parity+AFB <sup>a</sup> | 0.128 | -0.027 | 0.283 | 0.106 | 0.018 | -0.127 | 0.163 | 0.808 |
| +AAM+height+BMI18+Parity+AFB <sup>a</sup> | 0.116 | -0.040 | 0.273 | 0.146 | 0.007 | -0.140 | 0.154 | 0.923 |

AAM, Age at menarche; BMI, Body Mass Index at baseline, BMI18, BMI at age 18 years; AFB, Age at first birth.
Analyses adjusted for birth cohort (<1940, 1940-1949, 1950-1959, 1960-1969, 1970-1979, >=1980); and using weights derived as described in the Methods; analyses were also adjusted for age at interview
<sup>a</sup> only among parous women with information on AFB; AFB was considered as a categorical variable

**Table 4:** Interval between menopause and menarche and BRCA1 and BRCA2 PV carrier vs non-carrier status.

|  | **BRCA1 PV carriers** |  |  |  | **BRCA2 PV carriers** |  |  |  |
| --- | --- | --- | --- | --- | --- | --- | --- | --- |
|  | Estimate | L95CI | U95CI | p-value | Estimate | L95CI | U95CI | p-value |
| Birth cohort | -0.064 | -0.484 | 0.356 | 0.765 | -0.036 | -0.419 | 0.347 | 0.853 |
| +Parity+AFB+BMI<sup>a</sup> | -0.079 | -0.498 | 0.339 | 0.710 | -0.020 | -0.403 | 0.362 | 0.917 |
| +Parity+AFB+BMI18<sup>a</sup> | -0.025 | -0.445 | 0.394 | 0.905 | 0.001 | -0.384 | 0.387 | 0.994 |

AFB, age at first birth; BMI, Body Mass Index at baseline, BMI18, BMI at age 18 years
Linear regression analyses of interval between menopause and menarche in BRCA1 and BRCA2 PV carrier vs non-carriers were carried out using the norm.cens regression (VGAM); and using weights derived as described in the Methods.
<sup>a</sup> only among women with no missing information on AFB, parity or BMI, Parity and AFB treated as categorical variables

### Distribution of height and BMI among BRCA1 and BRCA2 carriers and non-carriers

BRCA1 PV carriers were slightly taller than non-carriers (mean difference 0.005m, p=0.003); for BRCA2 PV carriers the difference was 0.002m p=0.2 (Table 5). In unweighted analyses, the effect was also statistically significant for BRCA2 (p<0.05) (STable 6). The effect estimate was similar after adjusting for covariates BMI or BMI at age 18, AAM, height, parity, and age at first birth. There was no difference in BMI at age at interview or at age 18 years between carriers and non-carriers.

**Table 5:** Association between height, and BMI and BRCA1 and BRCA2 PV carrier vs non-carrier status.

| Trait | *BRCA1* PV carriers |  |  |  | *BRCA2* PV carriers |  |  |  |
| --- | --- | --- | --- | --- | --- | --- | --- | --- |
|  | estimate | L95CI | U95CI | p-value | estimate | L95CI | U95CI | p-value |
| height (m) |  |  |  |  |  |  |  |  |
| +Birth cohort (finer) | 0.005 | 0.002 | 0.009 | 0.003 | 0.002 | -0.001 | 0.006 | 0.212 |
| +Birth cohort (finer) +AAM+Parity+AFB <sup>a</sup> | 0.005 | 0.001 | 0.008 | 0.011 | 0.002 | -0.001 | 0.006 | 0.242 |
| +Birth cohort (finer) +BMI18+AAM+Parity+AFB <sup>a</sup> | 0.004 | 0.001 | 0.008 | 0.018 | 0.002 | -0.002 | 0.006 | 0.294 |
| BMI (kg) |  |  |  |  |  |  |  |  |
| +Birth cohort (finer) | -0.116 | -0.407 | 0.175 | 0.434 | 0.235 | -0.070 | 0.539 | 0.131 |
| +Birth cohort (finer)+AAM+height+Parity+AFB <sup>a</sup> | 0.011 | -0.274 | 0.295 | 0.940 | 0.272 | -0.025 | 0.569 | 0.072 |
| BMI at age 18 (kg) |  |  |  |  |  |  |  |  |
| +Birth cohort (finer) | -0.009 | -0.196 | 0.179 | 0.928 | 0.087 | -0.098 | 0.272 | 0.357 |
| +Birth cohort (finer)+AAM+height | 0.044 | -0.139 | 0.227 | 0.637 | 0.066 | -0.114 | 0.246 | 0.471 |

AAM, Age at menarche; BMI, BMI, Body Mass Index at baseline, BMI18, BMI at age 18 years; AFB, Age at first birth
all analyses adjusted for birth cohort (categorised as <1940, 1940-1949, 1950-1959, 1960-1969, 1970-1979, >=1980); and using weights derived as described in the Methods.
<sup>a</sup> Only women with no missing information on age at menarche, BMI, height and parity were included in the analyses; Parity and AFB treated as categorical variable

## Discussion

We compared the distributions of breast cancer risk factors including ANM, AAM, the interval between ANM and AAM, menstrual cycle length, height and BMI in a cohort of BRCA1 and BRCA2 PV carriers, and non-carriers, from a large national study.

We found no statistically significant differences in the distributions of any of these traits, apart from height. In unadjusted analyses among women reporting natural menopause, we observed a lower mean ANM in BRCA1 carriers compared with non-carriers. However, in naïve analyses not accounting for age at censoring, ANM will inevitably be lower in PV carriers, as natural menopause can only be observed if it takes place prior to RRSO. Analyses adjusting for age at censoring (the last age at which menopause could be observed) or allowing for censoring using the ‘norm.cens’ function in R corrected for this phenomenon, and we found no statistically significant difference in ANM when these analytical strategies were applied. The larger correction for BRCA1 PV carriers is consistent with the higher cancer risk and more frequent and earlier uptake of RRSO. We also conducted time-to-event analyses for ANM using Cox regression; these analyses also showed no association between ANM and PV status for either gene.

DNA damage and repair mechanisms are central in the biology of menopause and BRCA1 and BRCA2 proteins play a crucial role in the process of DNA double strand break repair through regulation of homologous recombination. It is therefore biologically plausible that these processes interact to influence ANM in carriers. Case-control analysis in UK Biobank data have reported earlier natural menopause in women harbouring PTVs in BRCA1 or BRCA2 (13). However, the number of carriers in that study were limited (N=32 BRCA1 and N=143 BRCA2 carriers). In addition, the effects were smaller in Ward et al (18), after removing women known to have undergone gynaecological surgeries. The same study (13), however, also reported an earlier ANM in carriers of PALB2 PTVs, an association that was replicated in data from the BRIDGES study (mean ANM difference 1.78 years) (38). Given the functional similarity between BRCA2 and PALB2, a similar effect on ANM might be expected, so this discrepancy is perplexing.

While only 14% and 21% of carriers experienced natural menopause in EMBRACE, our study included many more PV carriers than Ruth et al (13) and was sufficiently powered to detect differences at least of the magnitude estimated using UK Biobank data. For example, the 95%CI for the effect size in the linear regression would exclude a half year earlier (or later) mean ANM in both BRCA1 and BRCA2 PV carriers. Our results highlight that methodological considerations are important in studies to evaluate risk factors in PV carriers, particularly when evaluating the distribution of age at natural menopause. Interventions, including RRSO in PV carriers, complicate interpretation and results may be sensitive to measurement error. Menopause occurs over a period of time and the recording of both the timing and reason for menopause may be inaccurate. Recording of RRSO and cancer diagnoses may also be incomplete or inaccurate. A decision to undergo RRSO may be related to family history of ANM or cancer, as has been previously documented. Furthermore, RRSO may have been scheduled close to anticipated menopause.

There are limitations in the methodology used to assess associations with ANM. Linear regression ignores data on pre-menopausal women. On the other hand, the assumptions underlying Cox regression may not be met since the data arise from a cross-sectional study and are retrospective, rather than prospective, and censoring may be informative. It is also possible that recruitment might be influenced by menopausal status, but this seems unlikely since recruitment is largely determined by family history of cancer. While the effect size (hazard ratio for menopause associated with PV status) may not be a valid estimate of the effect size that would be obtained in a true prospective study, this should still provide a valid test of the difference in ANM by PV status. Moreover, the proportional hazards model underlying Cox regression may not be realistic. Modelling using the cens.norm function overcomes some of these issues, allowing for censoring whilst using all available data. Due to unbalanced sampling due to recruitment through genetics clinics, analyses with differential weighting of cases and controls were carried out. None of the above methods showed any evidence of association between PV status and ANM. Taken together they strongly suggest that any association is likely to be weak.

Another limitation is that non-carriers were only followed up until age at interview, and for this reason only information obtained via the baseline questionnaire was used for both carriers and non-carriers. Future studies providing accurate record linkage to surgeries and medication use, and more frequent follow-up to identify when women when first experience menopausal symptoms in relation to other life events, will be valuable.

Menarche, on the other hand, takes place well before the development of cancer, RRSO or genetic testing. We found no association between AAM and carrier status, though age at menarche may be inaccurately reported and could be susceptible to recall bias. BMI at baseline is likely to be accurately reported, and we found no difference in the distributions of BMI between carriers and non-carriers. We did, however, find a small but statistically significant difference in height between carriers and non-carriers, BRCA1 PV carriers being ∼0.5cm taller than non-carriers. Measurement of height is likely to be accurate and unbiased. Height is an established risk factor for breast cancer, and many of the biological pathways underlying growth are also relevant to cancer, but to our knowledge this has not so far implicated BRCA-related mechanisms. This observation could be a chance finding, and of note, the effect for BRCA2 PV carriers differs between the weighted and unweighted analyses, but if replicated it would be interesting to investigate the mechanisms underlying differences in height in carriers and implications for cancer risk.

A major strength of this study is comparability between carriers and non-carriers, many of whom are family members of carriers. On the other hand, it is possible that non-carriers are not entirely representative of the general population. Known and unknown factors relevant to membership of a PV carrier family, for example higher levels of screening, or healthy volunteer bias could be relevant.

In addition to the intrinsic biological interest, the results of these analyses have practical implications. The BOADICEA model assumes that the baseline distributions of risk factors are independent of genotype. If that were not the case, the model would need to be adapted to allow for genotype-specific distributions. These findings suggest that this should not be necessary, at least for the risk factors investigated in this study. In addition, under the assumption of risk-factor/genotype independence, it is possible to evaluate the interactions between risk factors in population-based studies using case-only analyses, which are more powerful than case-control analyses, particularly for rare exposures such as PV status. Currently, in the BOADICEA model lifestyle/hormonal risk factors are assumed to be associated with the same relative risk in PV carriers as non-carriers. It has proved difficult to obtain sufficient prospective data to evaluate this directly, and such case-only analyses may provide a more powerful basis to evaluate these interactions. This, in turn, should provide a reliable basis for counselling and management of PV carriers.

## Declarations

### Ethics approval and consent to participate

The EMBRACE study was approved by the East of England – Cambridge South Ethics Committee (ref 98/5/026, IRAS 20971). All participants gave written informed consent.

### Consent for publication

Not applicable

### Availability of data and materials

The datasets generated and/or analysed during the current study are not publicly available, as they potentially include personal data. However, they can be accessed upon reasonable request made to the EMBRACE study Data Access Coordination Committee and the completion of a data sharing agreement.

## Authors’ contributions

Writing Group: NM, ACA, DFE; Study design: NM, ACA, DFE; Data management: DF, EZ; Statistical Analysis: NM, DFE, DRB; Funding: ACA, DFE, MT, DGE, SA; Provided data: MA, JB, AFB, PB, HCon, JC, HCop, RD, AD, ED, DG, RH, LI, ZK, FL, ZM, PJM, JM, AM, HM, CS, LS, KS, VT, LW, SA, DGE, MT, ACA, DFE. All authors read and approved the final version of the manuscript.

## Acknowledgements

We thank all patients and clinicians participating in this study. A full list of EMBRACE Trusts and Principal investigators are included in STable 7.

## List of abbreviations

PV: Pathogenic variant
ANM: age at natural menopause
AAM: age at menarche
BMI: body mass index
RRSO: risk reducing salpingo-oophorectomy
EMBRACE: Epidemiological Study of Familial Breast Cancer
LOF: loss-of-function
ER: Oestrogen receptor
CI: Confidence intervals
p: p-value

